# Certified large language model-based diagnostic decision support in rheumatology: the ALLIANCE multicentre randomised controlled trial

**DOI:** 10.64898/2026.08.29.26361715

**Authors:** Phillip Kremer, Nadine Schlicker, Ragip Hasnaj, Jonathan Bamberger, Thorben Witte, Isabell Haase, Andreas Mayr, Carsten Schmidt, Nina Osteras, Xenofon Baraliakos, Sebastian Kuhn, Martin Krusche, Johannes Knitza

## Abstract

**Objectives:** To evaluate whether access to a certified large language model (LLM)-based clinical decision support system improves physician diagnostic performance in rheumatology compared with conventional diagnostic resources alone.

**Methods:** In this multicentre, open-label, randomised controlled trial, 82 physicians from seven hospitals in two countries were randomised 1:1 to conventional diagnostic resources plus Prof. Valmed or conventional resources alone. Participants assessed three rheumatology vignettes before and after assistance. The primary outcome was top-1 diagnostic accuracy. Secondary outcomes included top-3 accuracy, diagnostic reasoning, confidence, case-processing time and perceived support quality.

**Results:** Top-1 accuracy increased from 22.2% to 33.3% in the intervention group and from 23.3% to 35.0% in the control group, with no between-group difference in improvement (adjusted OR 0.99, 95% CI 0.45 to 2.19; p=0.979). Differences in top-3 accuracy, diagnostic reasoning and confidence were also not significant. Assisted case-processing time was substantially shorter with LLM support (94 vs 206 s; adjusted mean difference −112 s, 95% CI −141 to −83; p<0.001). Information timeliness and perceived diagnostic support quality were rated significantly higher in the intervention group. Exploratory analyses showed persistent overconfidence and substantial AI over-reliance.

**Conclusions:** Certified LLM-based diagnostic support did not improve diagnostic accuracy compared with conventional resources, but substantially reduced case-processing time and improved perceived support quality. These findings suggest potential workflow benefits while highlighting overconfidence and over-reliance as important safety considerations.

**Trial registration number:** NCT07166692

**WHAT IS ALREADY KNOWN ON THIS TOPIC:**

- Accurate and timely diagnosis in rheumatology is challenging because workforce shortages coincide with non-specific, complex and often rare presentations.
- Large language models are increasingly used for diagnostic decision support and have shown strong performance in benchmarking studies.
- Randomised evidence for physician-facing diagnostic decision support, particularly in rheumatology and for certified LLM-based systems, is lacking.

**WHAT THIS STUDY ADDS:**

- The ALLIANCE trial is the first randomised controlled trial to evaluate physician-facing diagnostic decision support in rheumatology and the first to assess a medically certified LLM-based clinical decision support system in any clinical specialty.
- A certified LLM-based clinical decision support system did not improve top-1 diagnostic accuracy compared with conventional resources alone, as assistance led to similar gains in both groups.
- Use of a certified LLM-based clinical decision support system was associated with approximately half the assisted case-processing time of conventional resources alone, as well as higher perceived support quality.

**HOW THIS STUDY MIGHT AFFECT RESEARCH, PRACTICE OR POLICY:**

- LLM-based diagnostic decision support may be most useful for improving efficiency, support quality, and broadening differential diagnoses rather than increasing top-1 diagnostic accuracy.
- Safe clinical implementation will require careful attention to overconfidence, over-reliance, transparency, and trust.
- Further robust trials, ideally under real-world clinical conditions, are needed to define the role of clinical decision support in routine care.

## Introduction

Accurate and timely diagnosis is essential in rheumatology, where delayed or incorrect diagnosis can postpone treatment and contribute to irreversible organ damage. Diagnostic assessment is challenging because many rheumatic diseases present with non-specific, overlapping symptoms and include rare conditions. Consequently, diagnostic delay^1,2^ and misdiagnosis are common^3,4^.

Clinical decision support systems (CDSS) have been investigated for decades^5^, and early systems showed promise in rheumatology^6^. However, implementation in routine care has often been limited by time-consuming data entry, modest performance, and poor interactivity^7–9^. Large language models (LLMs) may overcome some of these barriers by enabling conversational, flexible, and efficient diagnostic support^10^. In rheumatology, generative LLMs such as ChatGPT have demonstrated diagnostic performance comparable to that of experienced rheumatologists^11^. Patients with rheumatic diseases are also increasingly using LLMs themselves, and survey data suggest broad support for physician use of these tools as clinical decision support^12^. In parallel, physicians, including rheumatologists, are increasingly exploring generative LLMs for diagnostic decision-making^13–15^.

Despite this growing interest, general-purpose LLMs are not certified as medical devices, hence, their use in clinical practice may be restricted. Prof. Valmed was developed to address this translational and regulatory gap. The manufacturer describes it as the first LLM-based CDSS in Europe to receive CE certification as a medical device^16^. Prof. Valmed uses retrieval-augmented generation to anchor its outputs in curated scientific literature and thereby reduce hallucinations^16^. In prior evaluations, its diagnostic accuracy and processing time were comparable to commonly used systems such as ChatGPT and OpenEvidence^17^.

However, evidence from randomised evaluations of LLM-based diagnostic support remains limited. To our knowledge, no randomised controlled trial has evaluated physician-facing diagnostic decision support in rheumatology or assessed a certified LLM-based clinical decision support system^18,19^. Furthermore, existing randomised studies of general-purpose LLMs for diagnostic support have reported mixed findings, with some demonstrating improved physician diagnostic performance and others showing no significant benefit^18–25^.

We therefore conducted the ALLIANCE trial, a multicenter randomised controlled trial evaluating whether access to a certified LLM-based diagnostic decision support system improves physician diagnostic performance in rheumatology compared with conventional diagnostic resources alone. Secondary objectives were to assess diagnostic reasoning, case-processing time, diagnostic confidence, and perceived support quality.

## Methods

### Study design

ALLIANCE was a randomised, multicenter, open-label, parallel-group trial with blinded outcome assessment (ClinicalTrials.gov identifier: NCT07166692; first posted 10 September 2025). The study compared diagnostic support using conventional resources alone with diagnostic support using conventional resources plus the Prof. Valmed interface.

### Participants

Currently practicing physicians with training in rheumatology, internal medicine, emergency medicine, family medicine, dermatology, or orthopedics were recruited from seven hospitals in two countries: University Medical Center Hamburg-Eppendorf, University Hospital Marburg, University Hospital Erlangen, Rheumazentrum Ruhrgebiet, Klinikum Fulda, Charité Berlin, and Diakonhjemmet Hospital Oslo. Recruitment was conducted locally at each participating site using a voluntary convenience sampling strategy. Local study collaborators approached eligible physicians directly and invited them to participate. All participants provided written informed consent before enrollment and randomization. No financial compensation was offered.

### Randomization and masking

Following written informed consent, participants were randomised 1:1 to the intervention or control group. The allocation sequence was generated by an independent statistician using a computer-based random-number generator with variable block sizes of 2 and 4. Enrolling investigators had no access to the allocation sequence. Because of the nature of the intervention, participants were not blinded to group assignment, whereas outcome assessment was blinded. Randomization was stratified according to current or previous clinical practice in a rheumatology service.

### Procedures

Participants were presented sequentially with three clinical vignettes representing Cogan syndrome, dermatomyositis, and familial Mediterranean fever (see supplementary file). These vignettes were based exclusively on anamnestic information and were randomly selected from our previous rheumatology diagnostic decision support benchmarking study^10^. All study sessions were conducted remotely on an individual basis under live supervision by the study coordinator, with participants sharing their screens throughout the assessment. For each vignette, participants first provided a primary diagnosis, rated their diagnostic confidence, and could optionally list up to two additional differential diagnoses without assistance. They then completed the same task with assistance. In both groups, permitted resources included conventional sources such as established medical websites, including UpToDate and AMBOSS Medical Knowledge, as well as textbooks, whereas use of LLMs was not permitted.

In the intervention group, participants additionally had access to Prof. Valmed. This is a subscription-based software platform for diagnostic and therapeutic clinical decision support. It is described by the manufacturer as the first certified LLM-based medical device in Europe^16^. The system is based on curated medical literature and uses retrieval-augmented generation (RAG) to retrieve verified medical information, mitigate hallucinations, and generate case-specific responses rather than relying solely on pretrained model knowledge. The Prof. Valmed web interface was used during the study period without local control over backend versioning. As the system was accessed through the live web interface, participants used the version available at the time of each study session, reflecting the latest deployed version provided by the manufacturer. The study coordinator ensured standardized input, including a standardized zero-shot prompt and case information (supplementary material). The model was instructed to return three candidate diagnoses, each accompanied by a probability estimate and a brief explanation (supplementary material). After completing all cases, participants filled out a brief evaluation questionnaire (supplementary material) and could provide optional comments on their experience and perception of AI-based clinical decision support. Throughout the study sessions, the study coordinator monitored adherence to the protocol by ensuring that only permitted resources were consulted.

The study was reviewed by the institutional review board of Philipps-Universität Marburg, Germany, and was deemed exempt from formal ethical approval (24-221-1 ANZ). The trial reporting followed the CONSORT-AI 2020 extension for clinical trials involving artificial intelligence interventions. A completed CONSORT-AI checklist is provided in the supplementary material.

### Outcomes

The primary endpoint was top-1 diagnostic accuracy after assistance, defined as the proportion of cases in which the primary diagnosis exactly matched the reference diagnosis.

Secondary endpoints were: (1) top-3 diagnostic accuracy after assistance, defined as the proportion of cases in which the reference diagnosis was included among the three proposed diagnoses; (2) a cumulative diagnostic reasoning score, with 2 points assigned to correct diagnoses and 1 point to plausible diagnoses^10,17,24^; (3) physician-rated post-assistance diagnostic confidence, measured on a 0–10 visual analogue scale; (4) time required for assisted case completion in seconds; and (5) perceived information timeliness and (6) perceived diagnostic support quality of the assistance, both measured on a 5-point Likert scale and derived from the DeLone and McLean Information Systems Success Model^26^ and the measurement dimensions summarized by Petter et al.^27^.

### Diagnostic accuracy assessment

Diagnostic suggestions were evaluated independently and in a blinded manner by three board-certified rheumatologists, each with at least 8 years of clinical experience in rheumatology. Using previously described approaches, diagnostic suggestions were classified as identical, plausible, or diagnostically different relative to the reference diagnosis^10,28,29^. For each case, ratings were performed by two rheumatologists, one from University Hospital Marburg and one from University Medical Center Hamburg-Eppendorf, who were assigned at random. Before the formal assessment, all raters jointly reviewed and discussed 10 pilot vignettes to harmonize the rating procedure. Inter-rater agreement was substantial (Cohen’s κ=0.799). Disagreements were resolved by the third rheumatologist.

### Other Measures

Additional exploratory analyses examined diagnostic changes after assistance, differential diagnosis list length, diagnostic calibration, and stand-alone Prof. Valmed performance in relation to physician performance before and after assistance. After case completion, participants in the intervention group completed 5 questionnaire items on acceptance, usability ^30^ and trust, each rated on a 7-point Likert scale (supplementary material). Participants in both groups could also provide optional comments on their study experience and views on AI-based diagnostic support. These responses were summarized descriptively as exploratory findings.

### Statistical analysis

The required sample size of 82 participants was determined a priori. The calculation was based on the lower bound of the 95% confidence interval for the effect estimate reported in a previous trial by Roemer et al^20^, using a two-sided significance level of 0.05 and 80% power in a classical Fisher’s exact test. Because each participant completed three vignettes, the repeated-measures structure, assuming a within-participant correlation of 0.5, was expected to further increase statistical power.

To account for the three vignette assessments per participant, prespecified inferential analyses were performed using mixed-effects models with a random intercept for participant. The primary outcome, top-1 diagnostic accuracy, was analyzed using a longitudinal (two timepoints, pre- and post-assistance) generalized linear mixed model for binary outcomes with a logit link function. The model compared intervention and control groups and adjusted for age, gender, and specialty category (rheumatology vs non-rheumatology). The confirmatory treatment effect (group x time interaction) is reported as an adjusted odds ratio with corresponding 95% confidence interval. As a sensitivity analysis, assisted case-processing time was log-transformed to better satisfy model assumptions. Top-3 diagnostic accuracy was analyzed using an analogous generalized linear mixed model. Continuous secondary outcomes, including diagnostic confidence and diagnostic reasoning score, were analyzed using linear mixed-effects models adjusted for baseline values and the same covariates. Time required for case completion was summarized using mean and SD and analyzed using a linear mixed-effects model. Associations between study group and perceived information timeliness and perceived diagnostic support quality were analyzed using linear regression models adjusted for age, gender, and specialty category. Effect estimates for continuous outcomes are reported as adjusted mean differences with corresponding 95% CIs.

Statistical significance was defined as a two-sided p value of less than 0.05. The primary outcome was considered confirmatory. Analyses of secondary and exploratory outcomes were considered exploratory and were not adjusted for multiple comparisons. Additional exploratory analyses were performed to characterize changes in diagnostic behavior after assistance. Differential diagnosis list length before and after assistance was summarized separately for each study group using median and IQR. Post-assistance between-group differences were assessed using the Mann–Whitney U test. Exploratory post hoc analyses also compared stand-alone Prof. Valmed diagnostic accuracy with physician diagnostic accuracy before and after assistance, including comparisons stratified by rheumatology background. Pairwise comparisons between stand-alone Prof. Valmed and physician subgroup top-1 accuracy were assessed using Fisher’s exact tests. Exploratory calibration analyses were also performed across study conditions. For each group, mean diagnostic accuracy, mean confidence and diagnostic calibration were calculated. Diagnostic calibration was quantified using the O−U index, defined as mean confidence minus mean observed accuracy, with values ranging from −1 (complete underconfidence) to +1 (complete overconfidence), and 0 indicating perfect calibration^31,32^. We also performed exploratory analyses of AI over-reliance and under-reliance for the top diagnosis in the intervention group^33^. Over-reliance was the proportion of incorrect AI suggestions that physicians accepted, resulting in an incorrect final decision. Under-reliance was the proportion of correct AI suggestions that physicians rejected, also resulting in an incorrect final decision. Questionnaire responses relating to Prof. Valmed and optional participant comments were summarized descriptively and are reported as exploratory findings. The main inferential models were fitted in R version 4.3.1 (R Foundation for Statistical Computing, Vienna, Austria), including the lme4 package for mixed-effects modeling. Calibration analyses and data visualization, including Sankey diagrams, were performed in Python 3.13.5 (Python Software Foundation, Wilmington, Delaware, USA) using pandas, NumPy and Matplotlib.

## Results

### Participant characteristics

Of 112 physicians screened, 82 (21 rheumatologists, 61 non-rheumatologists) were randomised between October 12, 2025 and February 1, 2026 and provided data for the primary analyses. 42 physicians were randomised to the intervention group and 40 physicians to the control group (Figure 1). Demographic characteristics (Table 1) were generally comparable between groups, except for more dermatologists in the control group and more endocrinologists in the intervention group. The majority of physicians self-reported their sex as female (52.4%), mean age (SD) was 32.5 (7.0) years and median clinical experience was 3.5 years. Most physicians reported prior use of LLMs for medical purposes (89.0%) and indicated that they would welcome certified AI-based support for automatic differential diagnosis (82.9%).

**Figure 1.**
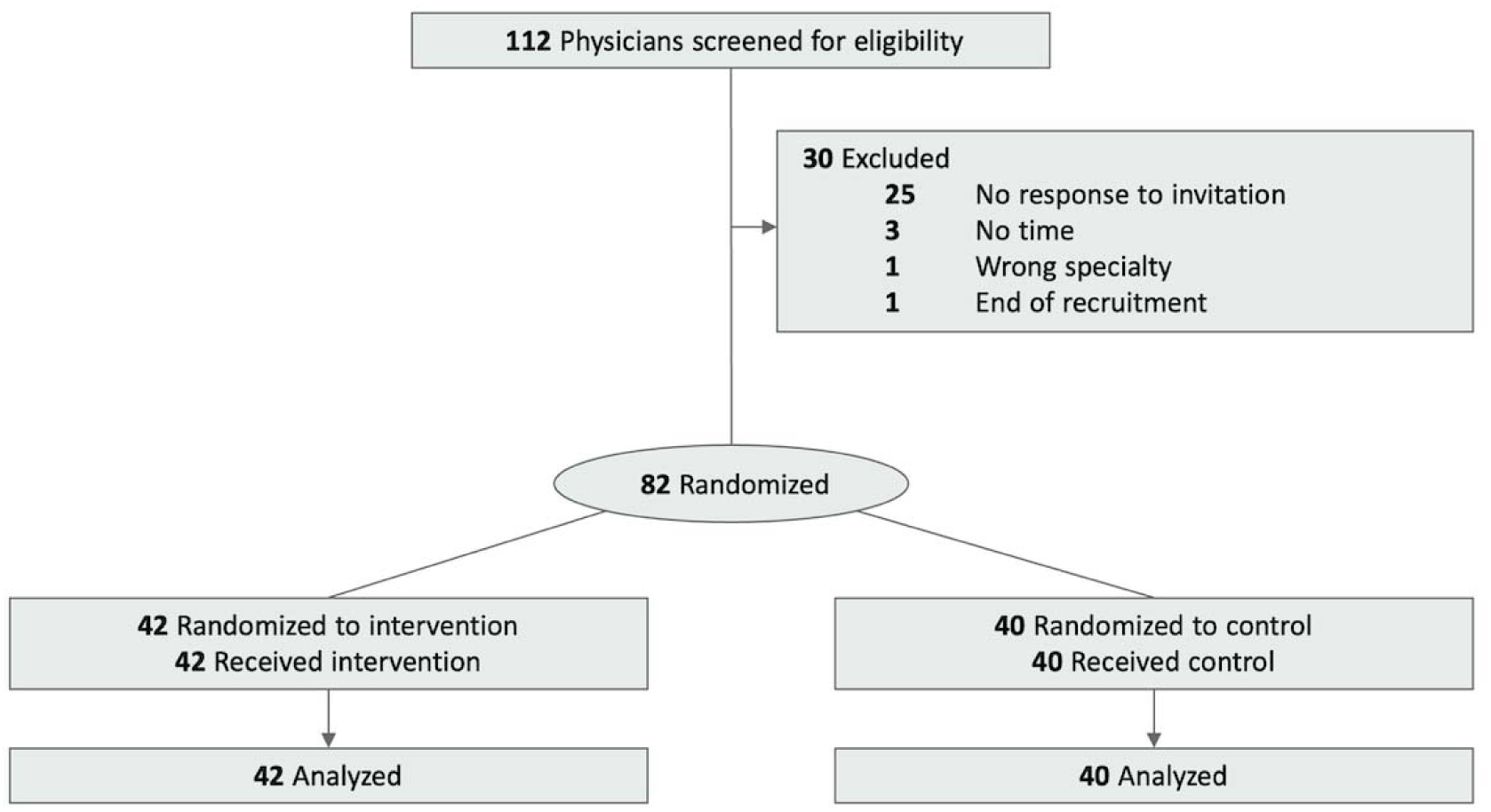
Trial Participant Flow Diagram.

**Table 1.** Baseline Characteristics (Intention-to-Treat Sample)

| Characteristic | Participants, No. (%) |  |  |
| --- | --- | --- | --- |
|  | Total (N = 82) | Intervention group (n = 42) | Control group (n = 40) |
| Age, mean (SD), y | 32.5 (7.0) | 33.2 (7.4) | 31.7 (6.6) |
| Clinical experience, median (IQR), y | 3.5 (2.0-6.0) | 4 (3.0-6.0) | 3 (1.4-6.0) |
| Previous LLM usage, n (%) |  |  |  |
| Previous medical LLM usage | 73 (89.0) | 38 (90.5) | 35 (87.5) |
| Only non-medical LLM usage | 8 (9.8) | 3 (7.1) | 5 (12.5) |
| No previous LLM usage | 1 (1.2) | 1 (2.4) | 0 (0) |
| Would welcome safe AI-based automatic differential diagnosis support, n (%) | 68 (82.9) | 34 (81.0) | 34 (85.0) |
| Gender, n (%) |  |  |  |
| Female | 43 (52.4) | 20 (47.6) | 23 (57.5) |
| Male | 39 (47.6) | 22 (52.4) | 17 (42.5) |
| Medical specialty, n (%) |  |  |  |
| Rheumatology | 21 (25.6) | 11 (26.2) | 10 (25.0) |
| Nephrology | 14 (17.1) | 7 (16.6) | 7 (17.5) |
| Cardiology | 8 (9.8) | 2 (4.8) | 6 (15.0) |
| Oncology | 2 (2.4) | 1 (2.4) | 1 (2.5) |
| Gastroenterology | 3 (3.7) | 1 (2.4) | 2 (5.0) |
| Endocrinology | 5 (6.1) | 5 (12.0) | 0 (0.0) |
| Pulmonology | 1 (1.2) | 0 (0.0) | 1 (2.5) |
| General internal medicine | 16 (19.5) | 11 (26.2) | 5 (12.5) |
| General medicine | 1 (1.2) | 1 (2.4) | 0 (0.0) |
| Dermatology | 5 (6.1) | 0 (0.0) | 5 (12.5) |
| Orthopedics | 4 (4.9) | 2 (4.8) | 2 (5.0) |
| Emergency medicine | 2 (2.4) | 1 (2.4) | 1 (2.5) |

### Primary outcome

Top-1 diagnostic accuracy before assistance was descriptively similar in the intervention and control groups (22.2% vs 23.3%). Post-assistance top-1 diagnostic accuracy was 33.3% in the intervention group and 35.0% in the control group, with no significant group-by-time interaction, indicating similar improvement from baseline in both groups (adjusted OR 0.99, 95% CI 0.45 to 2.19; p=0.979; Table 2; Figures 2 and 3).

**Figure 2.**
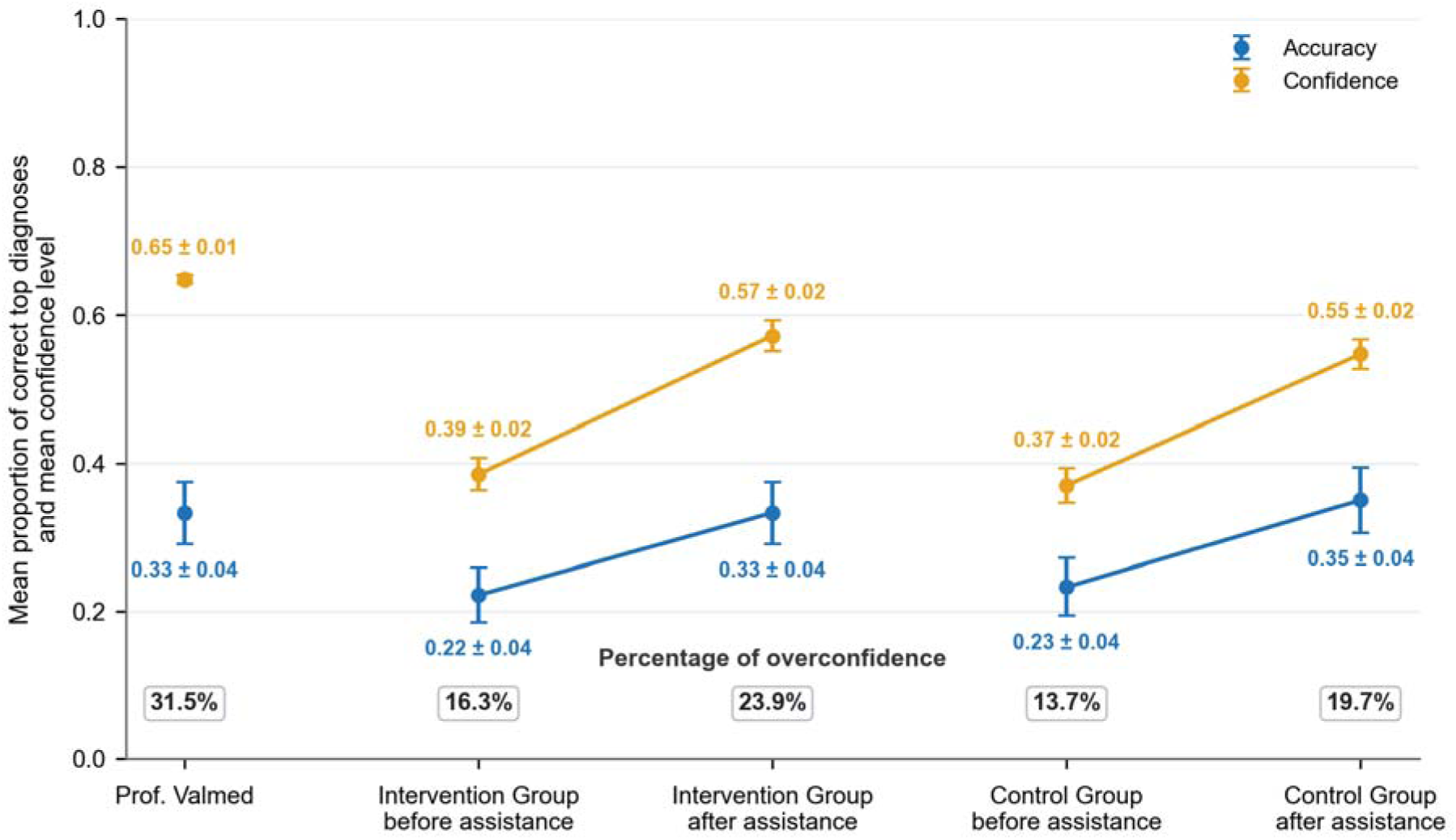
Diagnostic accuracy, confidence, and overconfidence across study conditions. Mean top-1 diagnostic accuracy and mean diagnostic confidence are shown for stand-alone Prof. Valmed, the intervention group before and after assistance, and the control group before and after assistance. Accuracy represents the proportion of correct top diagnoses, while confidence was rescaled to a 0–1 range. Points indicate mean values and error bars indicate standard errors. Numeric labels show mean ± standard error for accuracy and confidence. Calibration was defined as mean confidence minus mean accuracy, with values ranging from −1, indicating underconfidence, to +1, indicating overconfidence; values were rescaled and displayed below each condition as percentages. Overconfidence was observed across all settings and increased after assistance in both the intervention and control groups.

**Figure 3.**
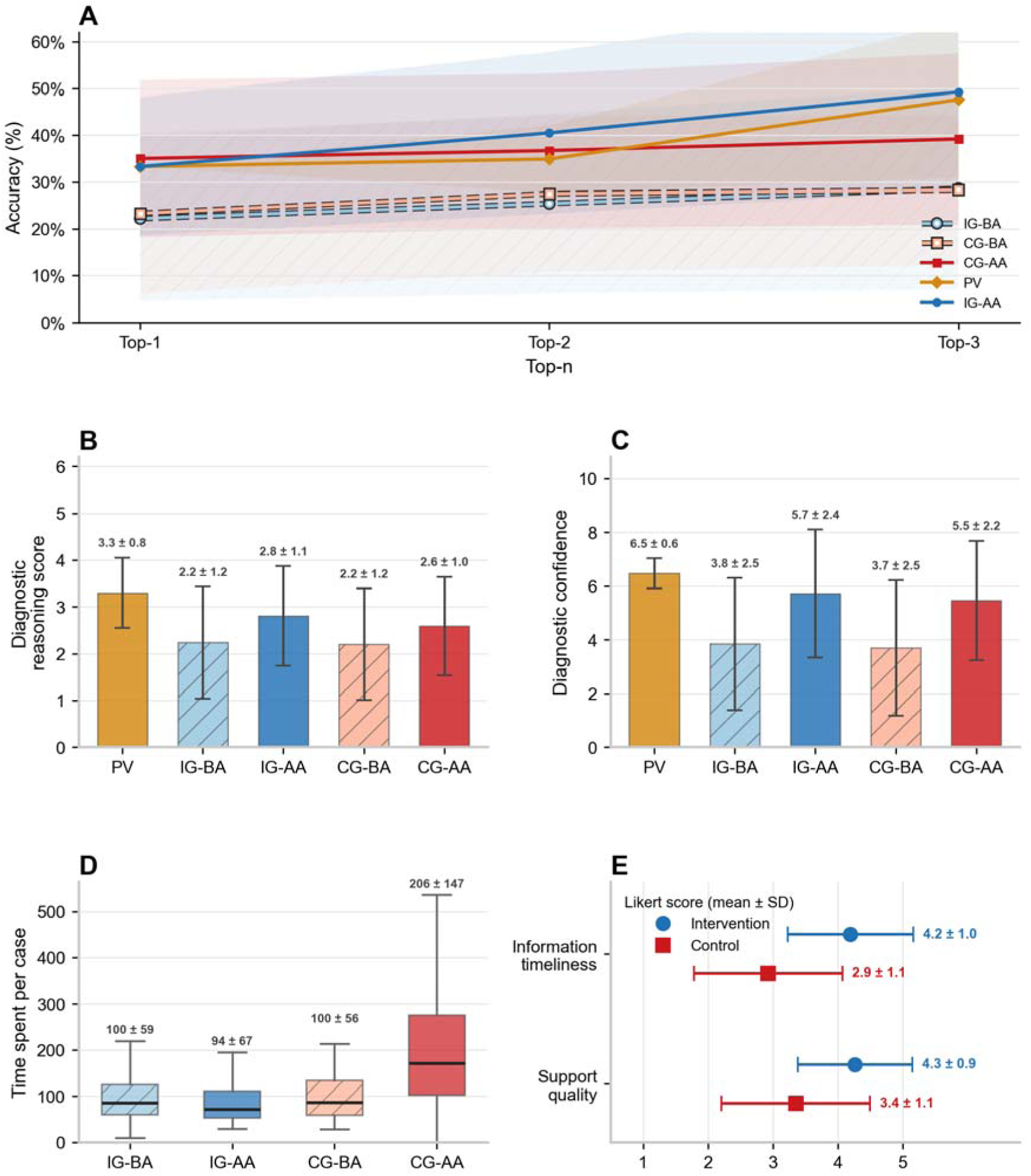
Diagnostic performance, confidence, time, and perceived support quality across study conditions. **A**, Top-n diagnostic accuracy before and after assistance in the intervention group and control group, with stand-alone Prof. Valmed performance shown for comparison. Accuracy is presented for top-1, top-2, and top-3 diagnostic suggestions. Shaded areas indicate mean and standard deviation bands. **B**, Diagnostic reasoning scores across stand-alone Prof. Valmed, intervention group before assistance, intervention group after assistance, control group before assistance, and control group after assistance. Bars show mean values and error bars indicate standard deviations. **C**, Diagnostic confidence across the same conditions, shown as mean ± SD. **D**, Time spent per case before and after assistance in the intervention and control groups. Boxes show the median and interquartile range, with whiskers indicating the non-outlier range. **E**, User-rated information support timeliness and overall perceived diagnostic support quality, measured on 5-point Likert scales. Points show mean values and error bars indicate standard deviations. Abbreviations: PV, Prof. Valmed stand-alone; IG-BA, intervention group before assistance; IG-AA, intervention group after assistance; CG-BA, control group before assistance; CG-AA, control group after assistance.

**Table 2.** Primary and secondary outcomes by study group, with exploratory stand-alone Prof. Valmed results.

| Outcome | Prof. Valmed stand-alone* | Intervention group before assistance | Intervention group post-assistance | Control group before assistance | Control group post-assistance | Effect estimate (95% CI) | p value |
| --- | --- | --- | --- | --- | --- | --- | --- |
| **Primary outcome** |  |  |  |  |  |  |  |
| Top-1 accuracy, n/N (%)<sup>†</sup> | 42/126 (33.3) | 28/126 (22.2) | 42/126 (33.3) | 28/120 (23.3) | 42/120 (35.0) | adjusted OR 0.99 (0.45 to 2.19) | 0.979 |
| **Secondary outcomes** |  |  |  |  |  |  |  |
| Top-3 accuracy, n/N (%)<sup>†</sup> | 60/126 (47.6) | 36/126 (28.6) | 62/126 (49.2) | 34/120 (28.3) | 47/120 (39.2) | adjusted OR 1.51 (0.56 to 1.79) | 0.291 |
| Diagnostic reasoning score, mean (SD)<sup>†</sup> | 3.3 (0.8) | 2.1 (1.2) | 2.8 (1.1) | 2.1 (1.2) | 2.6 (1.0) | adjusted mean difference 0.2 (-0.2 to 0.5) | 0.274 |
| Diagnostic confidence, mean (SD)<sup>†</sup> | 6.5 (0.6) | 3.8 (2.5) | 5.7 (2.4) | 3.7 (2.5) | 5.5 (2.2) | adjusted mean difference 0.1 (-0.6 to 0.8) | 0.760 |
| Time spent, mean (SD), s<sup>†</sup> | — | 100 (59) | 94 (67) | 100 (56) | 206 (147) | adjusted mean difference -112 (-141 to -83) | <0.001 |
| Perceived information timeliness, mean (SD)<sup>††</sup> | — | — | 4.2 (1.0) | — | 2.9 (1.1) | adjusted mean difference 1.31 (0.83 to 1.79) | <0.001 |
| Perceived diagnostic support quality, mean (SD)<sup>††</sup> | — | — | 4.3 (0.9) | — | 3.4 (1.1) | adjusted mean difference 0.93 (0.47 to 1.39) | <0.001 |

\* Stand-alone Prof. Valmed results are shown for exploratory contextual comparison. <sup>†</sup>Adjusted between-group comparisons; models were adjusted for gender, age, specialty and baseline values. <sup>††</sup>Adjusted between-group comparisons; models were adjusted for gender, age, and specialty.

### Secondary outcomes

Top-3 diagnostic accuracy before assistance was descriptively similar in the intervention and control groups (28.6% vs 28.3%). Post-assistance top-3 diagnostic accuracy was 49.2% in the intervention group and 39.2% in the control group, but the interaction effect was not significant, indicating similar improvement from baseline in both groups (adjusted OR 1.51, 95% CI 0.56 to 1.79; p=0.291; Table 2; Figure 3). We detected a similar pattern for the diagnostic reasoning score and diagnostic confidence. We descriptively observed similar values before assistance between intervention and control group (diagnostic reasoning: mean [SD], 2.1 [1.2] vs 2.1 [1.2]; diagnostic confidence: 3.8 [2.5] vs. 3.7 [2.5]), and higher values in the intervention group after assistance (diagnostic reasoning score: mean [SD], 2.8 [1.1] vs 2.6 [1.0]; diagnostic confidence: mean [SD], 5.7 [2.4] vs 5.5 [2.2]). We detected neither a significant interaction effect for the diagnostic reasoning score (adjusted mean difference 0.2, 95% CI −0.2 to 0.5; p=0.274) nor for diagnostic confidence (adjusted mean difference 0.1, 95% CI −0.6 to 0.8; p=0.760). Case-processing time showed a significant interaction effect (adjusted mean difference −112 s, 95% CI −141 to −83; p<0.001). Mean (SD) time before assistance was similar in the intervention and control groups (99.7 [58.6] s vs 100.2 [55.9] s), but after assistance was substantially lower in the intervention group (94 [67] s vs 206 [147] s). Perceived information timeliness was significantly higher in the intervention group than in the control group (mean [SD], 4.2 [1.0] vs 2.9 [1.1]); adjusted mean difference 1.31, 95% CI 0.83 to 1.79; p < .001). Perceived diagnostic support quality was likewise higher in the intervention group (mean [SD], 4.3 [0.9] vs 3.4 [1.1]); adjusted mean difference 0.93, 95% CI 0.47 to 1.39; p < .001).

### Additional exploratory analyses

#### Exploratory analyses by specialty and stand-alone tool performance

Figure 4 displays that, in exploratory post hoc analyses, stand-alone Prof. Valmed top-1 diagnostic accuracy was higher than physician accuracy before assistance. The difference was statistically significant compared with non-rheumatologists before assistance (33.3 vs 21.3%, p=0.025), but not compared with rheumatologists before assistance (33.3% vs 27.0%, p=0.409). After assistance, Prof. Valmed top-1 accuracy was similar to physician accuracy among both rheumatologists (33.3% vs 34.9%, p=0.871) and non-rheumatologists (33.3% vs 33.9%, p=1.000).

**Figure 4.**
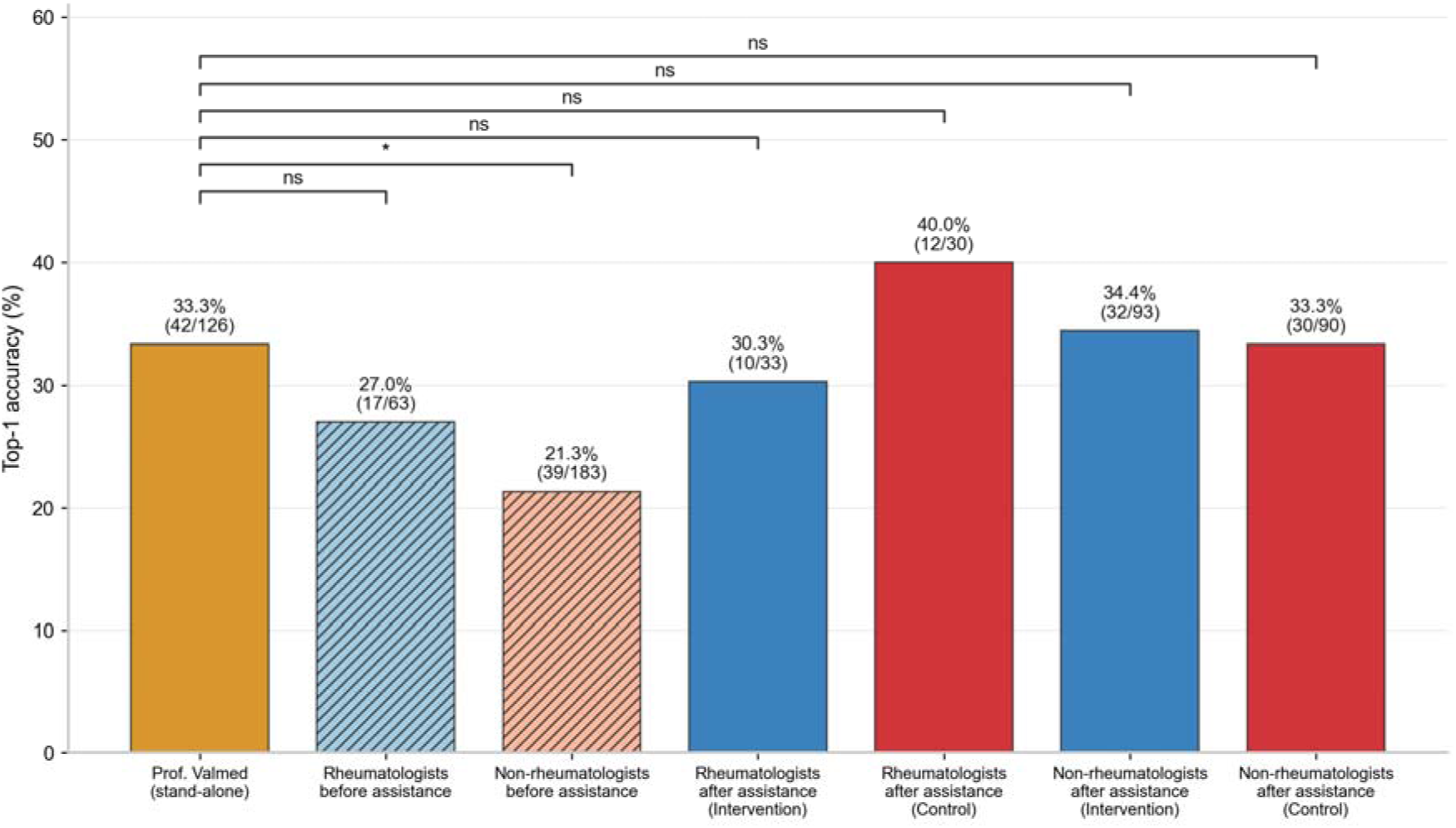
Stand-alone LLM-based CDSS performance compared with physician top-1 diagnostic accuracy by physician background and assistance condition. Top-1 accuracy is shown for stand-alone Prof. Valmed and for physicians stratified by rheumatology background before and after assistance. Bars indicate the proportion of cases with a correct top-1 diagnosis, with percentages and case counts shown above each bar. Before assistance, stand-alone Prof. Valmed achieved higher top-1 accuracy than both rheumatologists and non-rheumatologists, with a statistically significant difference compared with non-rheumatologists. After assistance, physician accuracy increased in both the intervention and control groups and was broadly similar to stand-alone Prof. Valmed performance. Brackets indicate pairwise comparisons with stand-alone Prof. Valmed; * denotes p<0.05 and ns denotes non-significant differences.

#### Effect of assistance

In both groups, the median differential diagnosis list length increased from 2 (IQR 1–3) before assistance to 3 (IQR 2–3) after assistance. Post-assistance list length did not differ significantly between groups (Mann–Whitney U test, p = 0.08). Among cases in which the top-1 diagnosis changed after assistance, accuracy improved in 25/29 cases (86.2%) and worsened in 4/29 cases (13.8%) in the intervention group; the corresponding proportions in the control group were 20/22 (90.9%) and 2/22 (9.1%), see Figure 5.

**Figure 5.**
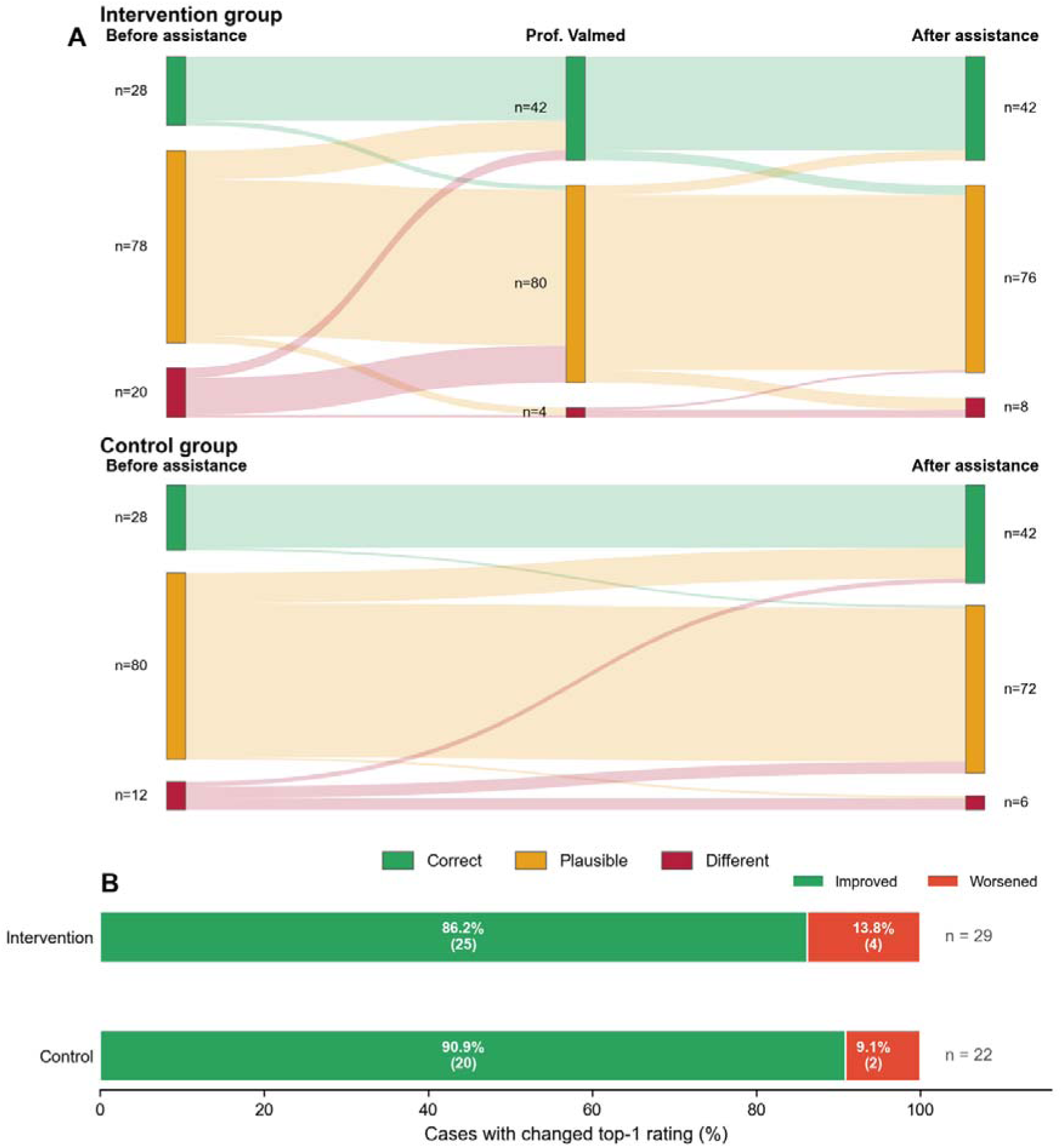
Changes in top-1 diagnostic ratings after assistance in the intervention and control groups. **A**, Sankey diagrams showing transitions in top-1 diagnostic ratings from before assistance to after assistance. In the intervention group, transitions are shown from physicians before assistance, through stand-alone Prof. Valmed output, to physicians after assistance. In the control group, transitions are shown from physicians before assistance to physicians after assistance. Diagnostic ratings were classified as correct, plausible, or different relative to the reference diagnosis. Flow widths represent the number of cases moving between rating categories. **B**, Among cases in which physicians changed their top-1 diagnosis after assistance, the proportion of changes that improved or worsened diagnostic accuracy is shown separately for the intervention and control groups. Most diagnostic changes improved accuracy in both groups, with similar proportions in the intervention and control groups.

#### Diagnostic calibration

Diagnostic calibration was characterized by overconfidence across all evaluated settings, peaking at 31.5% for stand-alone Prof. Valmed. In parallel with the increase in diagnostic confidence and diagnostic accuracy after assistance, overconfidence also increased in both groups, rising from 16.3% to 23.9% in the intervention group and from 13.7% to 19.7% in the control group, see Figure 2. In the intervention group, AI over-reliance was high (0.95), whereas AI under-reliance was low (0.10).

#### User acceptance in the intervention group

User acceptance of Prof. Valmed was high overall. Most participants reported that they would use the system again to support diagnosis in difficult cases (83%), found it easy to use (95%), and considered the interface pleasantly designed (86%). Trust in the system’s answers was somewhat lower (64%). By contrast, only 36% agreed that mistakes could be corrected quickly and easily, see Figure 6.

**Figure 6.**
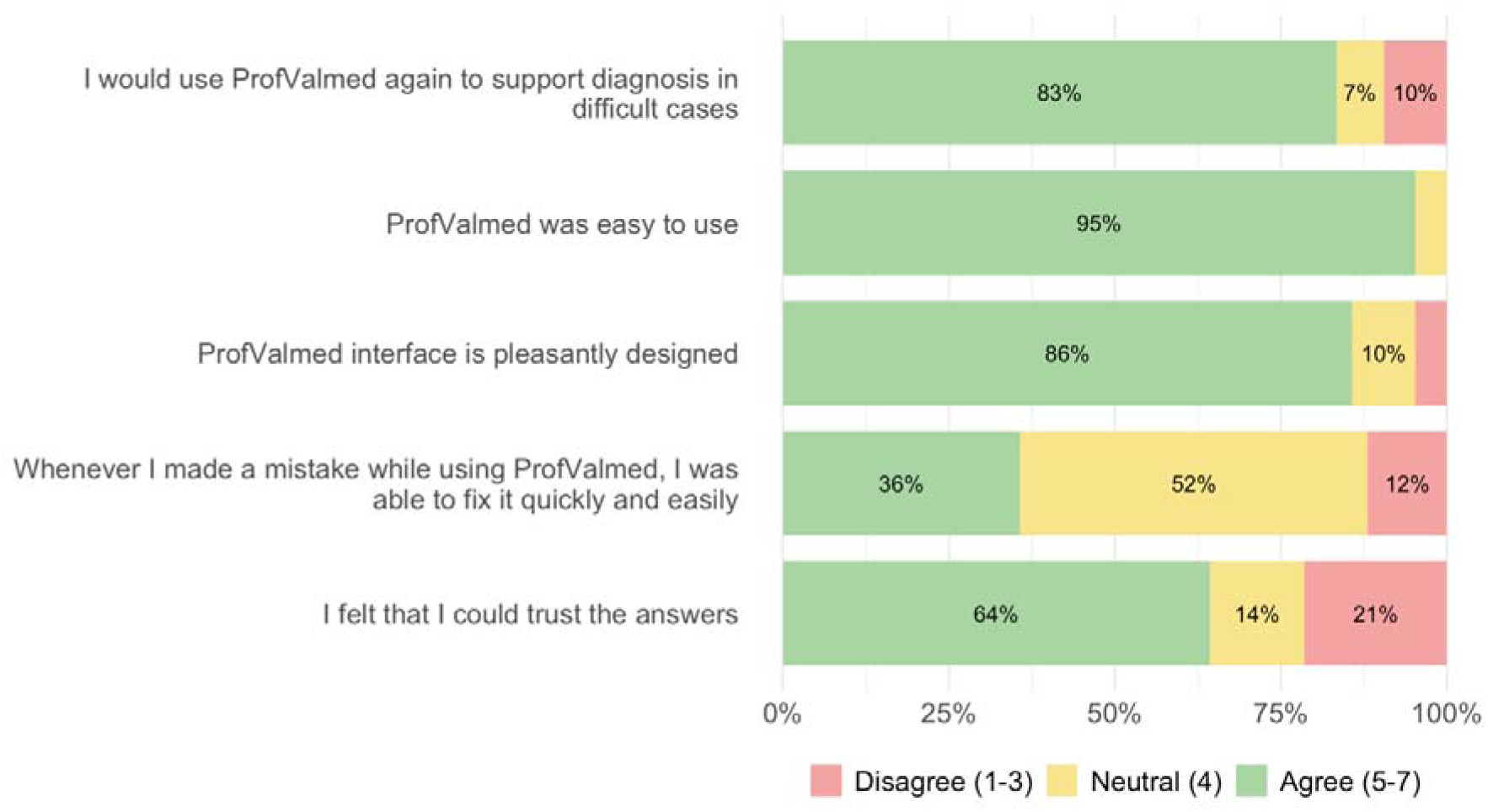
Physician-reported acceptance for Prof. Valmed. Stacked horizontal bars show the proportion of responses across three categories: disagree (scores 1–3), neutral (score 4), and agree (scores 5–7). Percentages are based on responses from 42 participants in the intervention group. User acceptance was high overall: most participants reported that they would use Prof. Valmed again to support diagnosis in difficult cases (83%), found it easy to use (95%), and considered the interface pleasantly designed (86%). Trust in the system’s answers was more moderate (64%), while fewer participants agreed that mistakes made while using Prof. Valmed could be corrected quickly and easily (36%).

#### Participant perspectives on AI-based diagnostic support

70 physicians provided optional free-text comments. The findings were broadly consistent with the high level of acceptance observed quantitatively for Prof. Valmed. Participants frequently described LLM-based support as easier, faster, and more practical for generating differential diagnoses than conventional resources such as UpToDate and AMBOSS, particularly when symptoms were vague. Several participants described informal use of generative AI tools, such as ChatGPT or OpenEvidence, in routine clinical practice outside formally governed institutional workflows, and indicated that they would welcome a secure, clinically integrated alternative. Participants particularly valued the ability of the LLMs to quickly generate differential diagnoses, connect disparate symptoms, and support reasoning through dialogue. At the same time, many emphasized that such tools should complement rather than replace physician judgment. Recurrent concerns included limited trust in the model output, unclear reasoning behind probability estimates, potential hallucinations, and the risk of overreliance. Participants also stressed the importance of transparent sources, data protection, and integration into existing clinical systems and workflows.

## Discussion

The ALLIANCE trial found that access to a certified LLM-based clinical decision support system did not improve top-1 diagnostic accuracy compared with conventional diagnostic resources alone. Top-1 accuracy increased to a similar extent in both groups after assistance, suggesting that both conventional and LLM-supported resources can support diagnostic performance. Beyond the primary outcome, LLM-based support was associated with numerically higher top-3 diagnostic accuracy, significantly shorter assisted case-processing time, and higher user-rated information timeliness and diagnostic support quality. In exploratory analyses, stand-alone Prof. Valmed achieved higher top-1 accuracy than unassisted non-rheumatologists and numerically higher accuracy than unassisted rheumatologists; however, after assistance, physicians in both study groups reached similar top-1 accuracy.

These results are clinically relevant because, in routine care, the value of diagnostic support may lie less in identifying a single leading diagnosis than in broadening the differential diagnosis, accelerating case assessment, and improving the perceived quality of diagnostic support. This interpretation is supported by the qualitative findings, in which participants frequently described LLM-based support as faster, easier and more practical than conventional tools for generating differential diagnoses, particularly in vague or atypical presentations. The shorter case-processing time observed with LLM support compared with conventional resources is consistent with recent studies^23,24^, although similar time savings have not been observed uniformly across all trials^20,21^. The absolute difference in our study was approximately 2 minutes per case, and the clinical relevance of such a gain is likely context dependent.

Our findings are consistent with the randomised trial by Goh et al.^24^, which likewise found no clear advantage of ChatGPT-assisted diagnosis over conventional resources. By contrast, other recent randomised trials have reported improved diagnostic performance with LLM support^21–23^. In two studies, stand-alone LLM performance exceeded unassisted physician performance, consistent with our exploratory findings for Prof. Valmed^21,22^. Notably, this contrasts with a previous direct rheumatologist-versus-LLM comparison in rheumatology, in which rheumatologists were numerically superior^11^. Whereas some earlier physician studies found that stand-alone LLM performance remained superior to physician performance with assistance^21,24^, our results align with more recent evidence suggesting that human–AI collaboration can achieve performance similar to AI alone^20,22,23^. Taken together, these findings align with the emerging literature and suggest that strong stand-alone model performance may improve physician accuracy relative to the unassisted setting^18,19^, without necessarily surpassing conventional support or the model alone^25^.

These results extend the evidence base beyond controlled benchmarking studies to a physician-facing, human-in-the-loop randomised evaluation. To our knowledge, this is the first randomised controlled trial to evaluate physician-facing diagnostic decision support in rheumatology and the first to assess a certified LLM-based clinical decision support system. The manufacturer-independent design, multicenter recruitment across two countries, and inclusion of both rheumatologists and non-rheumatologists further strengthen the study.

An interesting finding was that confidence exceeded observed accuracy across all evaluated settings and increased further after assistance in both groups, indicating persistent overconfidence. Exploratory analyses furthermore suggested substantial AI over-reliance in the intervention group, whereas under-reliance was uncommon. These findings suggest that diagnostic support may increase confidence more readily than correctness and highlight the need to evaluate calibration and behavioural reliance alongside accuracy. This concern is supported by a recent randomised trial in novice medical students^34^, in which misleading AI-generated explanations significantly reduced diagnostic accuracy, while correct explanations did not significantly improve accuracy over control. Misleading explanations also impaired confidence calibration and frequently induced commission errors, suggesting that plausible AI-generated misinformation can promote confident but incorrect decisions. This is also supported by evidence from other AI-assisted fields suggesting de-skilling with repeated use^35^, and prior rheumatology trial data demonstrating automation bias through acceptance of incorrect diagnostic suggestions^9^. Future studies should therefore examine not only whether AI-based decision support improves diagnostic accuracy, but also how it affects confidence calibration, commission errors, critical appraisal, over-reliance, and potential de-skilling.

The study has limitations. It was based on only three vignettes and relied exclusively on anamnestic information, limiting generalizability to real-world encounters that include examination findings, laboratory tests and imaging. All procedures were conducted remotely under supervised conditions. The use of Prof. Valmed was standardized through study-coordinator-mediated input, which improved consistency but may not reflect real-world interaction. In addition, recruitment followed a voluntary convenience sampling strategy, so selection bias cannot be excluded, particularly given the high baseline familiarity with LLMs in the sample. Several analyses, including subgroup comparisons, calibration analyses and stand-alone tool comparisons, were exploratory, based on small samples, and should be interpreted cautiously.

Future studies should evaluate LLM-based diagnostic support under routine clinical conditions, ideally with direct integration into clinical information systems and access to richer patient data. Such studies should examine not only diagnostic accuracy, but also downstream testing behavior, safety, calibration, workflow effects, and longer-term behavioral consequences.

In conclusion, ALLIANCE is, to our knowledge, the first randomised controlled trial of physician-facing diagnostic decision support in rheumatology. Use of a certified LLM-based clinical decision support system did not significantly improve top-1 diagnostic accuracy, top-3 diagnostic accuracy, or diagnostic reasoning compared with conventional diagnostic resources alone. However, it was associated with significantly shorter assisted case-processing time and higher user-rated information timeliness and support quality. Stand-alone LLM performance exceeded unassisted physician performance, while confidence exceeded accuracy across settings and increased after assistance. These findings suggest that LLM-based diagnostic support may be most valuable for improving efficiency, enhancing perceived support quality, and broadening differential diagnosis, while overconfidence and over-reliance remain key safety concerns. Further real-world trials are needed to define the role of certified LLM-based decision support in routine care.

## Data Availability

All data produced in the present study are available upon reasonable request to the authors

## Acknowledgements

We would like to thank all participants for their support of the study.

## Contributors

Concept and design: J.K., P.K., N.S., and A.M. Study coordination and investigation: P.K., R.H., T.W., I.H., C.S., N.O., X.B., S.K., M.K., and J.K. Data curation: P.K., N.S., J.B., A.M., and J.K. Data access and verification: P.K., N.S., A.M., and J.K. Formal analysis and visualization: J.B., N.S., P.K., A.M., and J.K. Methodology: P.K., N.S., A.M., S.K., M.K., and J.K. Supervision: C.S., S.K., X.B., M.K., and J.K. Drafting of the manuscript: J.K., N.S., and P.K. Critical revision for important intellectual content: all authors. All authors had full access to the data relevant to their contributions, approved the final version of the manuscript, and agreed to be accountable for all aspects of the work.

## Funding

Dr Knitza was supported in part by European Union Horizon grant 101080711 for SPIDERR. The funder had no role in study design, data collection, data analysis, data interpretation, manuscript preparation, or the decision to submit for publication. The manufacturer of Prof. Valmed did not fund the study and had no role in study design, data collection, data analysis, data interpretation, manuscript preparation, or the decision to submit for publication.

## Competing interests

The authors declare no financial or non-financial competing interests.

## Patient consent for publication

Not applicable

## Ethics approval

The study complies with the Declaration of Helsinki and was reviewed and deemed exempt from formal approval on 2 December 2024 by the institutional review board of Philipps-Universität Marburg, Germany (24-221-1 ANZ).

## Notes

### Competing Interest Statement

The authors have declared no competing interest.

### Clinical Trial

NCT07166692

### Author Declarations

Institutional review board of Philipps University Marburg Germany waived ethical approval for this work

## References

1 Benito-Lozano J, López-Villalba B, Arias-Merino G, et al. Diagnostic delay in rare diseases: data from the Spanish rare diseases patient registry. Orphanet J Rare Dis. 2022;17:418. doi: 10.1186/s13023-022-02530-3

2 Fuchs F, Morf H, Mohn J, et al. Diagnostic delay stages and pre-diagnostic treatment in patients with suspected rheumatic diseases before special care consultation: results of a multicenter-based study. Rheumatol Int. Published Online First: 10 October 2022. doi: 10.1007/s00296-022-05223-z

3 Gamez-Nava JI, Gonzalez-Lopez L, Davis P, et al. Referral and diagnosis of common rheumatic diseases by primary care physicians. British Journal of Rheumatology. 1998;37:1215–9. doi: 10.1093/rheumatology/37.11.1215

4 Knitza J, Tascilar K, Fuchs F, et al. Diagnostic Accuracy of a Mobile AI-Based Symptom Checker and a Web-Based Self-Referral Tool in Rheumatology: Multicenter Randomized Controlled Trial. J Med Internet Res. 2024;26:e55542. doi: 10.2196/55542

5 Miller RA, Pople HE, Myers JD. Internist-1, an experimental computer-based diagnostic consultant for general internal medicine. N Engl J Med. 1982;307:468–76. doi: 10.1056/NEJM198208193070803

6 Porter JF, Kingsland LC, Lindberg DA, et al. The AI/RHEUM knowledge-based computer consultant system in rheumatology. Performance in the diagnosis of 59 connective tissue disease patients from Japan. Arthritis Rheum. 1988;31:219–26. doi: 10.1002/art.1780310210

7 Abell B, Naicker S, Rodwell D, et al. Identifying barriers and facilitators to successful implementation of computerized clinical decision support systems in hospitals: a NASSS framework-informed scoping review. Implement Sci. 2023;18:32. doi: 10.1186/s13012-023-01287-y

8 Schwartz WB. Medicine and the computer. The promise and problems of change. N Engl J Med. 1970;283:1257–64. doi: 10.1056/NEJM197012032832305

9 Knitza J, Tascilar K, Gruber E, et al. Accuracy and usability of a diagnostic decision support system in the diagnosis of three representative rheumatic diseases: a randomized controlled trial among medical students. Arthritis Res Ther. 2021;23:233,. doi: 10.1186/s13075-021-02616-6.

10 Kremer P, Schiebisch H, Lechner F, et al. Comparative analysis of large language models and traditional diagnostic decision support systems for rare rheumatic disease identification. EULAR Rheumatology Open. 2025;1:51–9. doi: 10.1016/j.ero.2025.04.007

11 Krusche M, Callhoff J, Knitza J, et al. Diagnostic accuracy of a large language model in rheumatology: comparison of physician and ChatGPT-4. Rheumatol Int. Published Online First: 24 September 2023. doi: 10.1007/s00296-023-05464-6

12 Labinsky H, Klemm P, Graalmann L, et al. Patient experiences, attitudes, and profiles regarding artificial intelligence in rheumatology: a German national cross-sectional survey study. Rheumatol Int. 2025;45:269. doi: 10.1007/s00296-025-06023-x

13 Esnaashari S, Hashem Y, Francis J, et al. Exploring doctors’ perspectives on generative-AI and diagnostic-decision-support systems. BMJ Health Care Inform. 2025;32:e101371. doi: 10.1136/bmjhci-2024-101371

14 La Bella S, Rebollo-Giménez AI, Aouad K, et al. Artificial intelligence in rheumatology and paediatric rheumatology: insights from an international survey by EMEUNET. EULAR Rheumatology Open. 2026;100153. doi: 10.1016/j.ero.2026.03.001

15 Holzer M-T, Meinecke A, Müller F, et al. Artificial intelligence in rheumatology: status quo and quo vadis-results of a national survey among German rheumatologists. Ther Adv Musculoskelet Dis. 2024;16:1759720X241275818. doi: 10.1177/1759720X241275818

16 Medical co-pilot: First AI-supported medical device to support clinical decisions. 2025. https://www.vde.com/en/press/press-releases/medical-co-pilot-vde-aiq-valmed (accessed 28 April 2026)

17 Kremer P, Langballe E, Haase I, et al. Diagnostic performance of Prof. Valmed, ChatGPT-5 Thinking, and OpenEvidence in rheumatology: A comparative evaluation. Rheumatol Int. 2026;46:31. doi: 10.1007/s00296-025-06068-y

18 Chen SF, Alyakin A, Seas A, et al. LLM-assisted systematic review of large language models in clinical medicine. Nat Med. 2026;32:1152–9. doi: 10.1038/s41591-026-04229-5

19 Tretow I, Schwebel M, Feuerriegel S, et al. The Effect of LLM Assistance on Diagnostic Accuracy: A Meta-Analysis. 2025;2025.12.11.25341476.

20 Roemer A, Schlicker N, Kernder A, et al. Large language models enhance diagnostic reasoning of medical students in rheumatology: a randomized controlled trial. BMC Med Educ. Published Online First: 25 March 2026. doi: 10.1186/s12909-026-09079-w

21 McDuff D, Schaekermann M, Tu T, et al. Towards accurate differential diagnosis with large language models. Nature. Published Online First: 9 April 2025. doi: 10.1038/s41586-025-08869-4

22 Everett SS, Bunning BJ, Jain P, et al. From tool to teammate in a randomized controlled trial of clinician-AI collaborative workflows for diagnosis. npj Digit Med. Published Online First: 18 March 2026. doi: 10.1038/s41746-026-02545-1

23 Goh E, Bunning B, Khoong EC, et al. Physician clinical decision modification and bias assessment in a randomized controlled trial of AI assistance. Commun Med. 2025;5:59. doi: 10.1038/s43856-025-00781-2

24 Goh E, Gallo R, Hom J, et al. Large Language Model Influence on Diagnostic Reasoning: A Randomized Clinical Trial. JAMA Netw Open. 2024;7:e2440969. doi: 10.1001/jamanetworkopen.2024.40969

25 Vaccaro M, Almaatouq A, Malone T. When combinations of humans and AI are useful: A systematic review and meta-analysis. Nat Hum Behav. 2024;8:2293–303. doi: 10.1038/s41562-024-02024-1

26 Urbach N, Müller B. The Updated DeLone and McLean Model of Information Systems Success. In: Dwivedi YK, Wade MR, Schneberger SL, eds. Information Systems Theory: Explaining and Predicting Our Digital Society, Vol. 1. New York, NY: Springer 2012:1–18.

27 Petter S, DeLone W, McLean E. Measuring information systems success: models, dimensions, measures, and interrelationships. Eur J Inf Syst. 2008;17:236–63. doi: 10.1057/ejis.2008.15

28 Hautz WE, Kämmer JE, Hautz SC, et al. Diagnostic error increases mortality and length of hospital stay in patients presenting through the emergency room. Scand J Trauma Resusc Emerg Med. 2019;27:54. doi: 10.1186/s13049-019-0629-z

29 Bastakoti M, Muhailan M, Nassar A, et al. Discrepancy between emergency department admission diagnosis and hospital discharge diagnosis and its impact on length of stay, up-triage to the intensive care unit, and mortality. Diagnosis. 2022;9:107–14. doi: 10.1515/dx-2021-0001

30 Zimmermann J, Morf H, Scharf F, et al. German Version of the Telehealth Usability Questionnaire and Derived Short Questionnaires for Usability and Perceived Usefulness in Health Care Assessment in Telehealth and Digital Therapeutics: Instrument Validation Study. JMIR Hum Factors. 2024;11:e57771. doi: 10.2196/57771

31 Meyer AND, Payne VL, Meeks DW, et al. Physicians’ Diagnostic Accuracy, Confidence, and Resource Requests: A Vignette Study. JAMA Intern Med. 2013;173:1952. doi: 10.1001/jamainternmed.2013.10081

32 Weber N, Brewer N. Confidence-accuracy calibration in absolute and relative face recognition judgments. J Exp Psychol Appl. 2004;10:156–72. doi: 10.1037/1076-898X.10.3.156

33 Ma S, Wang X, Lei Y, et al. “Are You Really Sure?” Understanding the Effects of Human Self-Confidence Calibration in AI-Assisted Decision Making. Proceedings of the CHI Conference on Human Factors in Computing Systems. Honolulu HI USA: ACM 2024:1–20.

34 Teng D, Tan L, Cao Q, et al. Impact of AI misinformation on diagnostic accuracy and confidence calibration in novice medical students. npj Digit Med. Published Online First: 17 March 2026. doi: 10.1038/s41746-026-02547-z

35 Budzyń K, Romańczyk M, Kitala D, et al. Endoscopist deskilling risk after exposure to artificial intelligence in colonoscopy: a multicentre, observational study. Lancet Gastroenterol Hepatol. 2025;10:896–903. doi: 10.1016/S2468-1253(25)00133-5

