## Supplementary file 1 for "Certified large language model-based diagnostic decision support in rheumatology: the ALLIANCE multicentre randomised controlled trial"

**Supplementary material**

### 1. Prompt, including respective case descriptions

**Case vignette 1: Cogan syndrome**

“Based on the following patient information, provide a list of up to 3 possible diseases that the patient might have and short reasons why. The list should be sorted by probability from most to least probable. Assign each disease a probability to indicate the likelihood of the diagnosis given the anamnesis. The estimated probabilities should represent certainty and do not need to sum up to 100%. Patient Information: *61-year-old man. Presented with tinnitus, progressive hearing loss, generalized joint pain, blurred vision, and redness in both eyes.*
Possible diseases and probabilities in percent:”

Original source^1^

Standardized vignette^2^

**Case vignette 2: Dermatomyositis**

“Based on the following patient information, provide a list of up to 3 possible diseases that the patient might have and short reasons why. The list should be sorted by probability from most to least probable. Assign each disease a probability to indicate the likelihood of the diagnosis given the anamnesis. The estimated probabilities should represent certainty and do not need to sum up to 100%. Patient Information: *47-year old male with 3-day history of swelling around his left eye and a sensation of tightness in his throat. He was hoarse but did not have wheezing or shortness of breath. During the 3 days before presentation, it had become difficult for him to swallow solids, and he felt as if food was sticking in his throat. The patient was allergic to shellfish but had no known recent exposure. He took no prescription medications and had not used any new over-the-counter medications in the preceding month*. Possible diseases and probabilities in percent:”

Original source^3^

Standardized vignette^2^

**Case vignette 3: Familial mediterranean fever**

“Based on the following patient information, provide a list of up to 3 possible diseases that the patient might have and short reasons why. The list should be sorted by probability from most to least probable. Assign each disease a probability to indicate the likelihood of the diagnosis given the anamnesis. The estimated probabilities should represent certainty and do not need to sum up to 100%. Patient Information: *22-year old male from Azerbaijan with recurrent fever lasting 3 to 4 days since 1 year and abdominal pain.*
Possible diseases and probabilities in percent:“

Original source: University Hospital Hamburg-Eppendorf

Standardized vignette^2^

**Vignette references**

1.	Chen, L., Teng, J., Yang, C. & Chi, H. Cogan syndrome following SARS-COV-2 infection. *Clin Rheumatol* **42**, 2517–2518 (2023).

2.	Kremer, P. *et al.* Comparative analysis of large language models and traditional diagnostic decision support systems for rare rheumatic disease identification. *EULAR Rheumatology Open* **1**, 51–59 (2025).

3.	MacFarlane, L., Osman, N., Ritter, S., Miller, A. L. & Loscalzo, J. CLINICAL PROBLEM-SOLVING. Eye of the Beholder. *N Engl J Med* **374**, 1774–1779 (2016).

### 2. Evaluation questionnaire

**All participants**

**1.** I was able to receive the information needed without delay *****

I absolutely disagree	1 ( )	2 ( )	3 ( )	4 ( )	5 ( )	I absolutely agree

**2.** The diagnostic support was helpful *****

I absolutely disagree	1 ( )	2 ( )	3 ( )	4 ( )	5 ( )	I absolutely agree

**3.** Please share any thoughts on your assistance experience and perception of AI-based clinical decision support (free text):

**Intervention group only**

**1.** I would use ProfValmed again to support diagnosis in difficult cases.

I absolutely disagree	1 ( )	2 ( )	3 ( )	4 ( )	5 ( )	6 ( )	7 ( )	I absolutely agree

**2.** ProfValmed was easy to use.

I absolutely disagree	1 ( )	2 ( )	3 ( )	4 ( )	5 ( )	6 ( )	7 ( )	I absolutely agree

**3.** ProfValmed interface is pleasantly designed.

I absolutely disagree	1 ( )	2 ( )	3 ( )	4 ( )	5 ( )	6 ( )	7 ( )	I absolutely agree

**4.** Whenever I made a mistake while using ProfValmed, I was able to fix it quickly and easily.

I absolutely disagree	1 ( )	2 ( )	3 ( )	4 ( )	5 ( )	6 ( )	7 ( )	I absolutely agree

**5.** I felt that I could trust the answers. *****

I absolutely disagree	1 ( )	2 ( )	3 ( )	4 ( )	5 ( )	6 ( )	7 ( )	I absolutely agree

### 3. CONSORT-AI Extension Checklist

Adapted from Liu X, Cruz Rivera S, Moher D, Calvert MJ, Denniston AK, and the SPIRIT-AI and CONSORT-AI Working Group. Reporting guidelines for clinical trial reports for interventions involving artificial intelligence: the CONSORT-AI extension. Nature Medicine 26, 1364-1374 (2020), Table 1. Licensed under CC BY 4.0. Use alongside the CONSORT 2010 Explanation and Elaboration.

| **Section** | **CONSORT 2010 item** | **CONSORT-AI item** | **Addressed on page number** |
| --- | --- | --- | --- |
| **Title and abstract** | | | |
| Title and abstract | **1a.** Identification as a randomized trial in the title  **1b.** Structured summary of trial design, methods, results, and conclusions (for specific guidance see CONSORT for abstracts) | **CONSORT-AI 1a,b - Elaboration** (i) Indicate that the intervention involves artificial intelligence/machine learning in the title and/or abstract and specify the type of model.  (ii) State the intended use of the AI intervention within the trial in the title and/or abstract. | 1  2-3 |
| **Introduction** | | | |
| Background and objectives | **2a.** Scientific background and explanation of rationale  **2b.** Specific objectives or hypotheses | **CONSORT-AI 2a (i) - Extension** Explain the intended use of the AI intervention in the context of the clinical pathway, including its purpose and its intended users (for example, healthcare professionals, patients, public). | 6-7  7 |
| **Methods** | | | |
| Trial design | **3a.** Description of trial design (such as parallel, factorial) including allocation ratio  **3b.** Important changes to methods after trial commencement (such as eligibility criteria), with reasons |  | 7  - |
| Participants | **4a.** Eligibility criteria for participants  **4b.** Settings and locations where the data were collected | **CONSORT-AI 4a (i) - Elaboration** State the inclusion and exclusion criteria at the level of participants.  **CONSORT-AI 4a (ii) - Extension** State the inclusion and exclusion criteria at the level of the input data.  **CONSORT-AI 4b - Extension** Describe how the AI intervention was integrated into the trial setting, including any onsite or offsite requirements. | 7  8  8-9 |
| Interventions | **5.** The interventions for each group with sufficient details to allow replication, including how and when they were actually administered | **CONSORT-AI 5 (i) - Extension** State which version of the AI algorithm was used.  **CONSORT-AI 5 (ii) - Extension** Describe how the input data were acquired and selected for the AI intervention.  **CONSORT-AI 5 (iii) - Extension** Describe how poor quality or unavailable input data were assessed and handled.  **CONSORT-AI 5 (iv) - Extension** Specify whether there was human-AI interaction in the handling of the input data, and what level of expertise was required of users.  **CONSORT-AI 5 (v) - Extension** Specify the output of the AI intervention.  **CONSORT-AI 5 (vi) - Extension** Explain how the AI intervention’s outputs contributed to decision-making or other elements of clinical practice. | 9  8-9  -  8-9  9  8-9 |
| Outcomes | **6a.** Completely defined pre-specified primary and secondary outcome measures, including how and when they were assessed  **6b.** Any changes to trial outcomes after the trial commenced, with reasons |  | 9-10  - |
| Sample size | **7a.** How sample size was determined  **7b.** When applicable, explanation of any interim analyses and stopping guidelines |  | 11  - |
| Randomization: sequence generation | **8a.** Method used to generate the random allocation sequence  **8b.** Type of randomization; details of any restriction (such as blocking and block size) |  | 8  8 |
| Randomization: allocation concealment mechanism | **9.** Mechanism used to implement the random allocation sequence (such as sequentially numbered containers), describing any steps taken to conceal the sequence until interventions were assigned |  | 8 |
| Randomization: implementation | **10.** Who generated the random allocation sequence, who enrolled participants, and who assigned participants to interventions |  | 8 |
| Blinding | **11a.** If done, who was blinded after assignment to interventions (for example, participants, care providers, those assessing outcomes) and how  **11b.** If relevant, description of the similarity of interventions |  | 8 |
| Statistical methods | **12a.** Statistical methods used to compare groups for primary and secondary outcomes  **12b.** Methods for additional analyses, such as subgroup analyses and adjusted analyses |  | 11-12  12-13 |
| **Results** | | | |
| Participant flow (a diagram is strongly recommended) | **13a.** For each group, the numbers of participants who were randomly assigned, received intended treatment, and were analyzed for the primary outcome  **13b.** For each group, losses and exclusions after randomization, together with reasons |  | 27, Figure 1  27, Figure 1 |
| Recruitment | **14a.** Dates defining the periods of recruitment and follow-up  **14b.** Why the trial ended or was stopped |  | 13  - |
| Baseline data | **15.** A table showing baseline demographic and clinical characteristics for each group |  | 25 |
| Numbers analyzed | **16.** For each group, number of participants (denominator) included in each analysis and whether the analysis was by original assigned groups |  | 26 |
| Outcomes and estimation | **17a.** For each primary and secondary outcome, results for each group, and the estimated effect size and its precision (such as 95% confidence interval)  **17b.** For binary outcomes, presentation of both absolute and relative effect sizes is recommended |  | 26  26 |
| Ancillary analyses | **18.** Results of any other analyses performed, including subgroup analyses and adjusted analyses, distinguishing pre-specified from exploratory |  | 15-16 |
| Harms | **19.** All important harms or unintended effects in each group (for specific guidance see CONSORT for harms) | **CONSORT-AI 19 - Extension** Describe results of any analysis of performance errors and how errors were identified, where applicable. If no such analysis was planned or done, justify why not. | 10, 12-15 |
| **Discussion** | | | |
| Limitations | **20.** Trial limitations, addressing sources of potential bias, imprecision, and, if relevant, multiplicity of analyses |  | 19 |
| Generalizability | **21.** Generalizability (external validity, applicability) of the trial findings |  | 19 |
| Interpretation | **22.** Interpretation consistent with results, balancing benefits and harms, and considering other relevant evidence |  | 17-20 |
| **Other information** | | | |
| Registration | **23.** Registration number and name of trial registry |  | 7 |
| Protocol | **24.** Where the full trial protocol can be accessed, if available |  | - |
| Funding | **25.** Sources of funding and other support (such as supply of drugs), role of funders | **CONSORT-AI 25 - Extension** State whether and how the AI intervention and/or its code can be accessed, including any restrictions to access or re-use. | 9, 21 |
